# Risk and determinants of tuberculosis recurrence: a 12-year population-based cohort study

**DOI:** 10.1101/2025.11.20.25340642

**Authors:** E. Lepka de Lima, I. Salles, S. Cohn, A.A. Lindoso, S. Fukasava, J.E. Golub, S.R. Cox

**Author notes:** **Corresponding author:** Evelyn Lepka De Lima, MD, MPH. Technical Coordination, Municipal Tuberculosis Control Program, Municipal Health Secretariat of Guarujá, Av. Santos Dumont, 800 – Vila Santo Antônio, Guarujá, SP, Brazil, 11432-502.; **Alternate corresponding author**: Samyra R. Cox, PhD, MPH. Assistant Scientist, Department of International Health, Global Disease Epidemiology & Control Program, Bloomberg School of Public Health, Johns Hopkins University, 615 North Wolfe Street, Baltimore, MD 21205, USA.

## Abstract

**Background:** Tuberculosis recurrence remains a challenge to elimination efforts, particularly in high-burden settings. A better understanding of incidence, timing, and risk factors is needed to inform policies and programs for post-treatment care.

**Methods:** We conducted a retrospective cohort study of tuberculosis survivors successfully treated for a first tuberculosis episode between 2013 and 2024 in São Paulo state, Brazil. Data were obtained from the state tuberculosis surveillance system (TBweb), and deaths were identified through linkage with the national mortality database. We applied competing risks survival analysis to estimate incidence and assess risk factors for recurrence. Covariates included demographic, clinical, behavioral, and social vulnerability indicators, all measured at treatment initiation for the first tuberculosis episode. Risk factor models were stratified by age (<15 vs. ≥15 years).

**Findings:** Among 154,579 individuals who completed treatment for a first episode of tuberculosis, we identified 9,464 first recurrences over a median follow-up of 5.4 years (IQR 2.5–8.2). The overall recurrence incidence was 1,127.2 per 100,000 person-years. Rates were highest between 3–12 months after treatment completion, yet recurrence incidence remained over 1,000 per 100,000 person-years in the 2–5 years after treatment completion. In adults, hospitalization (subdistribution hazard ratio [SHR] 2.53, 95%CI 2.36–2.71), and incarceration during the initial episode (2.30, 2.13–2.48), together with pulmonary tuberculosis as initial diagnosis (2.10, 1.87–2.35) were the strongest independent risk factors for recurrence. In children, HIV (6.38, 2.64–15.40) was the strongest predictor.

**Interpretation:** The high and persistent risk of tuberculosis recurrence reinforces the importance of post-treatment care with risk-based follow-up strategies.

## Introduction

Over 155 million people worldwide are tuberculosis survivors.^1^ These survivors confront chronic pulmonary complications, reduced health-related quality of life, and heightened risk of future tuberculosis episodes compared to those without prior disease. ^2^This increased susceptibility is accompanied by elevated all-cause mortality and reduced life expectancy, regardless of successful initial treatment. ^3^

According to the World Health Organization (WHO), recurrent tuberculosis occurs when individuals who have successfully completed treatment experience another episode of tuberculosis. ^4^ This recurrence can arise from relapse (reactivation of the same infection) or exogenous reinfection. ^4^ Recurrent tuberculosis constitutes 5–20% of the global tuberculosis burden, with higher proportions observed in high-prevalence settings and among people infected with HIV. ^5–7^

Brazil has the highest tuberculosis burden in the Americas and is one of the WHO’s priority countries for tuberculosis elimination. ^8^ Recurrence poses a significant challenge to tuberculosis control in the country and other high-burden settings. Recent studies found 6.5-7.1% of people diagnosed with tuberculosis in Brazil were experiencing recurrence, which was linked to unfavorable outcomes such as loss of follow-up, treatment failure, and death. ^9,10^

Still, significant gaps remain in understanding who is most at risk for recurrence, when it occurs, and what factors drive it. The limited evidence on post-treatment outcomes has hindered development of clear guidance on post-treatment care, such as optimal duration of follow-up, identification of subgroups requiring closer surveillance, and appropriate structure of post-treatment monitoring. Methodological strategies, such as the use of large datasets, extended follow-up periods, and consideration of competing risks, are essential for robust characterization of tuberculosis recurrence dynamics. These approaches allow for more precise estimates of recurrence incidence and help identify key risk factors, thereby informing post-treatment monitoring, particularly in high-burden settings where structured follow-up can have the most significant impact. We therefore utilized Brazil’s well-established surveillance databases to conduct a large-scale retrospective cohort study across São Paulo state to estimate the incidence and predictors of recurrent tuberculosis and address these evidence gaps.

## Methods

### Study design and population

We constructed our cohort using data from the São Paulo State Tuberculosis Program, including all tuberculosis notifications recorded in the TBweb system from January 1, 2013, to December 31, 2024. TBWeb is an electronic platform implemented in 2006 to register and monitor all individuals diagnosed with tuberculosis across São Paulo state. It allows real-time data entry and management across all municipalities and is integrated with SINAN, the national database for notifiable diseases in Brazil. Each individual is assigned a unique SINAN number, enabling longitudinal follow-up, linking multiple tuberculosis episodes for the same person, and avoiding record duplication. Data were extracted from TBweb in January 2025. São Paulo is the most populous Brazilian state (44 million inhabitants) ^11^, reporting nearly one-quarter of national tuberculosis cases, and an incidence of 42.6 per 100,000 population. ^12^

For this study, individuals with a first tuberculosis episode with recorded treatment completion (i.e., were at risk for a first tuberculosis recurrence) were considered eligible. We included only those with a treatment outcome of ‘cure’ in their first tuberculosis episode, as reported in the TBWeb system — an administrative classification based on clinical, radiologic, and/or microbiological criteria. Among those, we included only cases whose treatment duration— the period between treatment initiation and closure recorded in the system — was within the expected range based on national guidelines. ^13^ Specifically, those with less than five months or greater than 2.5 years of treatment were excluded. Individuals treated for five to six months were included only if they had two consecutive negative smear microscopy results at months five and six. Individuals not classified as “cured” were excluded regardless of treatment duration. For the first tuberculosis episode, we also excluded individuals reclassified as non-tuberculosis, those who died during treatment, those without a recorded outcome, and those without a clearly identifiable first tuberculosis episode during the observation period.

Finally, we excluded records with missing outcome dates, irreparable inconsistencies, or duplications. For individuals with multiple notifications beyond the first recurrence (e.g., retreatment entries or additional recurrence episodes), only data and follow-up time up to the first recurrence were retained; subsequent notifications were excluded.

We obtained official written permission to use data from the data guardian (São Paulo State Health Department) and ethics approval from the Research Ethics Committee of the Instituto de Infectologia Emílio Ribas, São Paulo, Brazil, and the Institutional Review Board at Johns Hopkins University School of Medicine.

### Data abstraction and definition of potential risk factors

Data were abstracted from the TBweb system. Potential risk factors were selected *a priori* for risk factor analysis based on biological plausibility, relevance to tuberculosis recurrence described in literature, and data quality. All variables were measured at baseline, corresponding to the time of first tuberculosis diagnosis. No longitudinal updates of comorbidity status or treatment information were available in TBWeb. Demographic factors included age at tuberculosis diagnosis, sex assigned at birth (female or male), self-reported race (White, Black, Brown [“Pardo” – admixed Black and White], Asian, and Indigenous, according to the Brazilian Institute of Geography and Statistics – IBGE), and education level (classified as None; 1 – 7; 8 – 11; 12 or more years of schooling). Clinical vulnerability factors comprised comorbidities such as HIV, diabetes, mental illness, and other non-HIV immunosuppressive conditions (defined by clinical diagnosis or self-report as documented in TBWeb), as well as disease-related characteristics such as the form of tuberculosis (pulmonary, extrapulmonary, or both), hospitalization during the first episode, and mode of treatment administration (supervised or self-administered). Laboratory confirmation of tuberculosis was defined as a positive smear microscopy, culture, or molecular test result. Health-related behaviors included current alcohol, tobacco, and illicit drug use. Indicators of social vulnerability included experiencing homelessness or incarceration, all assessed at the time of first tuberculosis diagnosis and treatment initiation.

### Outcome Measurement

Our primary outcome was the first tuberculosis recurrence episode, defined as a new tuberculosis episode occurring after documented treatment completion for the initial tuberculosis episode. This definition is consistent with the WHO definition ^4^, which does not specify a minimum time interval between episodes. Individuals entered the cohort at the date of treatment completion for their first tuberculosis episode and were followed until first recurrence, death, or administrative censoring on December 31, 2024.

The first tuberculosis recurrence was assessed only among individuals with the initial and subsequent tuberculosis episodes documented in the system to ensure accurate sequencing of events. For each recurrence, the event date was defined as the date of treatment initiation.

Individuals with multiple recurrences contributed only their first recurrence to the analysis. Death was considered a competing event and identified through probabilistic linkage with the Brazilian Mortality Information System (SIM) to obtain date and cause of death. Details of the linkage process are provided in the appendix.

### Statistical Analysis

We described baseline characteristics by recurrence status. Categorical variables were summarized as counts and percentages and compared using the chi-squared test. As a continuous variable, age was summarized as mean and standard deviation (SD). Survival analysis used time from first tuberculosis treatment completion to recurrence, death, or censoring.

Cumulative incidence functions (CIFs) were estimated to assess the probability of first tuberculosis recurrence over time and compared using Gray’s test. ^14^ Because death during follow-up precludes recurrence and represents a competing risk, we used a Fine and Gray model to estimate the cumulative incidence of recurrence and evaluate associated risk factors. Results were expressed as subdistribution hazard ratios (SHRs) with 95% confidence intervals (CIs). All potential confounders were included simultaneously in multivariable models without automated selection procedures. Analyses were stratified by age at first tuberculosis diagnosis (<15 years and ≥15 years), with separate models for each group. For individuals ≥15 years, models included sociodemographic, clinical, behavioral, and social vulnerability variables. For individuals <15 years, a more restricted model was applied, excluding education, incarceration, homelessness, and substance use, as these variables were not consistently assessed in children. Among the included variables, collinearity was assessed using Pearson correlation and variance inflation factors (VIF), with a cutoff of 5.

Our primary survival analyses used a complete-case approach. To assess the robustness of our findings, we conducted a sensitivity analysis using multiple imputation by chained equations (MICE), under the assumption of missing at random (MAR). Ten imputed datasets were generated including all model covariates, the event indicator, and follow-up time.

Competing-risk models were rerun, and results were pooled using Rubin’s rules.

All statistical tests were two-tailed, with a significance level of 0.05. Analyses were conducted using Stata (version 19.5; StataCorp, College Station, TX, USA) and R (version 4.5.0; R Foundation for Statistical Computing, Vienna, Austria).

## Results

Of 249,908 tuberculosis notifications recorded between January 2013 and December 2024, 95,329 were excluded based on predefined criteria (Figure 1). These included individuals not at risk for first tuberculosis recurrence (53,764), those with insufficient information for follow-up (14,380), those with invalid treatment timelines (6,420), and subsequent notifications related to retreatment or recurrences beyond the first event, not eligible for analysis (20,765).

**Figure 1.**
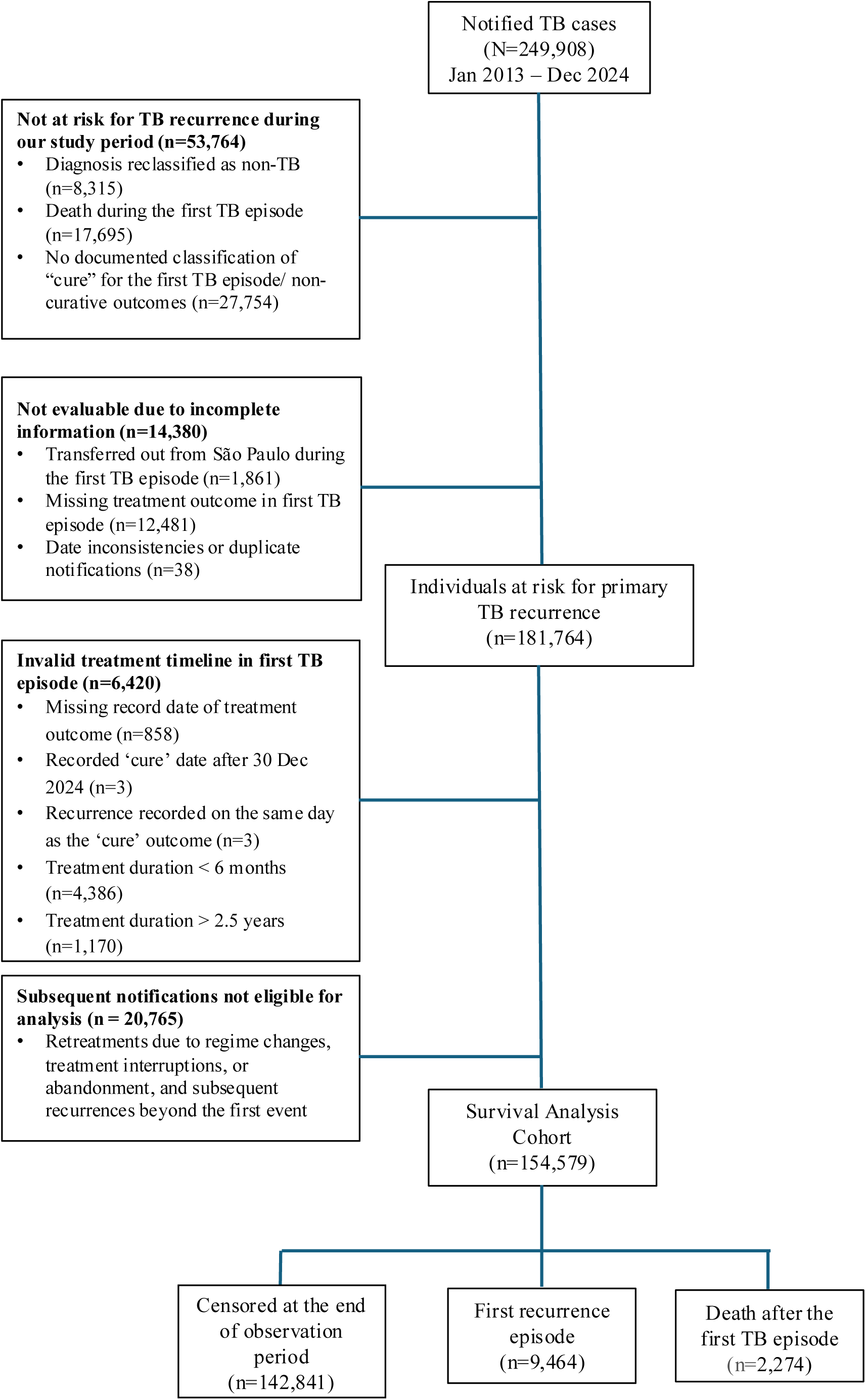
Selection of the analytic cohort for first tuberculosis recurrence analysis Among 249,908 tuberculosis notifications recorded in São Paulo (2013–2024), cases were excluded in four categories: (1) not at risk for tuberculosis recurrence; (2) not evaluable due to incomplete information; (3) invalid treatment timelines; and (4) subsequent notifications not eligible for time-to-first recurrence analysis. See Supplementary Table 1 for detailed criteria and case counts.

The final analytical cohort comprised 154,579 tuberculosis survivors who were at risk for recurrent tuberculosis. At the time of the first tuberculosis diagnosis, participants had a mean age of 37.7 years (SD 16.4), and the majority were male (107,646; 69.6%). Most individuals self-identified as Black or Brown (65,789; 52.5%), and the predominant clinical form was pulmonary tuberculosis (126,793; 82.0%). Laboratory confirmation was documented for 120,259 (77.8%) individuals. HIV infection and diabetes were reported in 9,409 (6.5%) and 10,741 (6.9%) participants, respectively. Incarceration and homelessness during first diagnosis were recorded in 20,074 (13.4%) and 3,367 (2.3%) of participants. Reported substance use included alcohol (23,638; 15.8%), illicit drugs (21,947; 14.7%), and tobacco (33,237; 22.3%). Most individuals (117,237; 81.4%) received supervised treatment (Table 1).

**Table 1.**
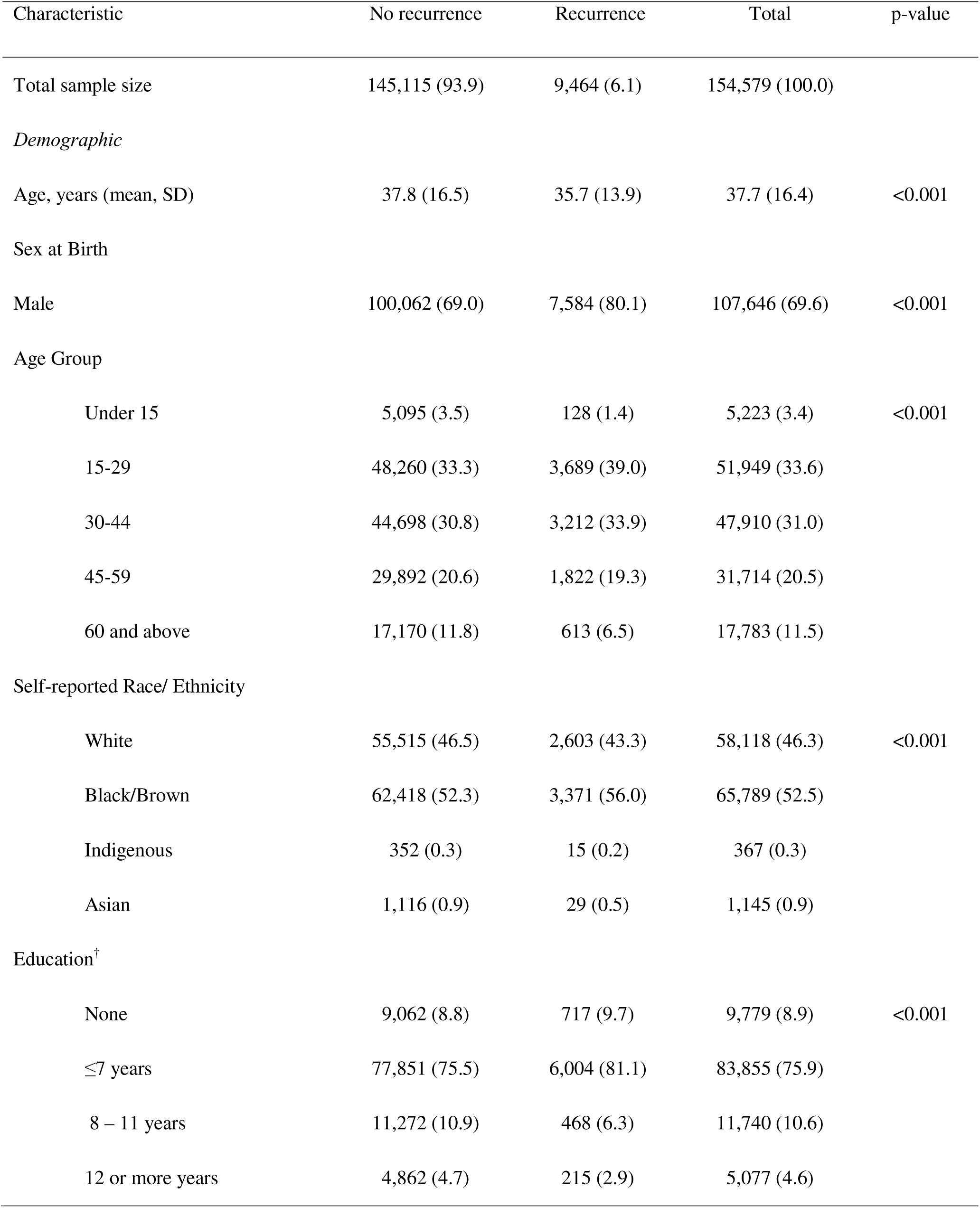
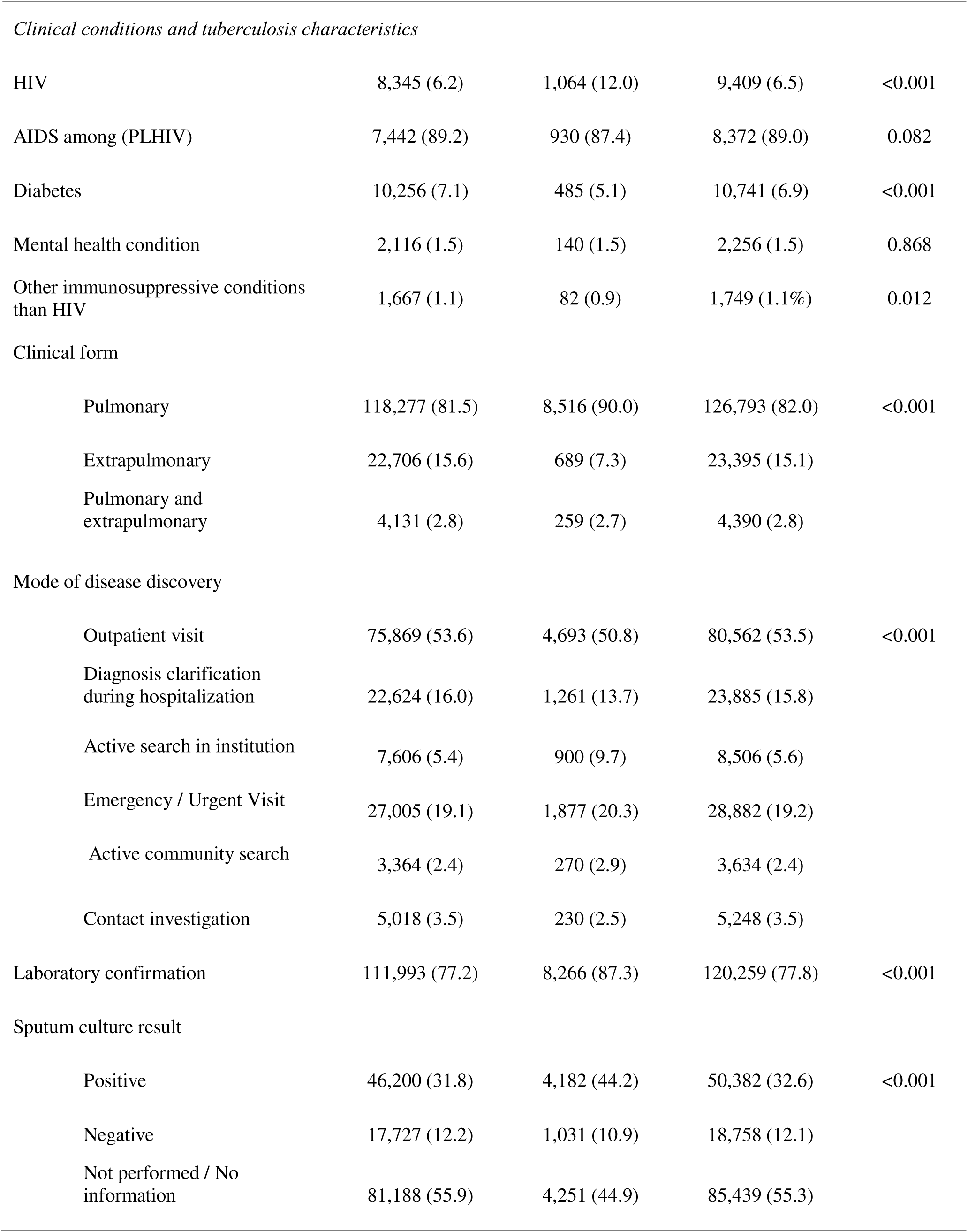
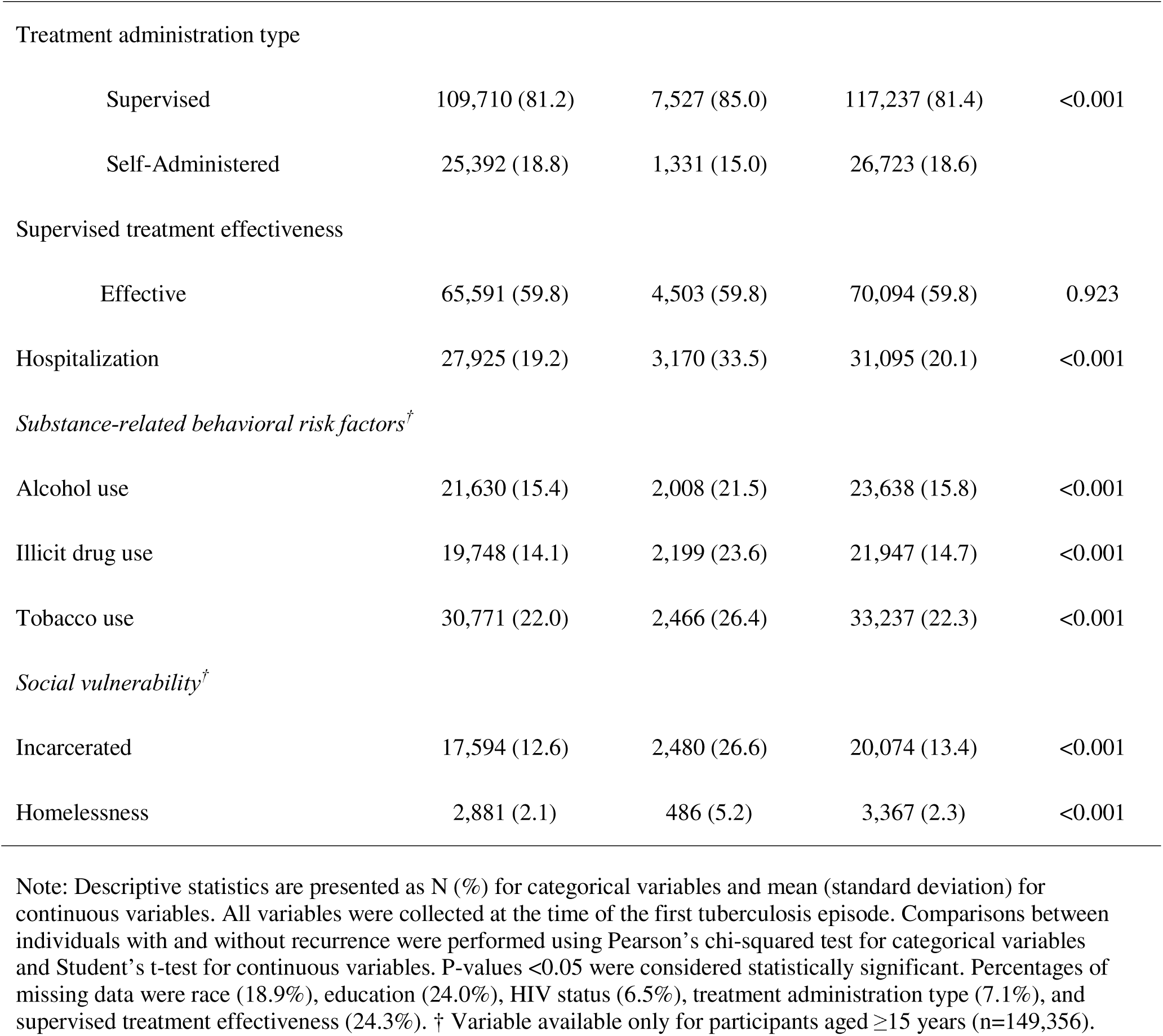
Baseline characteristics of individuals who completed treatment for a first episode of tuberculosis and were included in the recurrence risk analysis, São Paulo, Brazil (2013–2024)

| Characteristic | No recurrence | Recurrence | Total | p-value |
| --- | --- | --- | --- | --- |
| Total sample size | 145,115 (93.9) | 9,464 (6.1) | 154,579 (100.0) |  |
| *Demographic* |  |  |  |  |
| Age, years (mean, SD) | 37.8 (16.5) | 35.7 (13.9) | 37.7 (16.4) | <0.001 |
| Sex at Birth |  |  |  |  |
| Male | 100,062 (69.0) | 7,584 (80.1) | 107,646 (69.6) | <0.001 |
| Age Group |  |  |  |  |
| Under 15 | 5,095 (3.5) | 128 (1.4) | 5,223 (3.4) | <0.001 |
| 15-29 | 48,260 (33.3) | 3,689 (39.0) | 51,949 (33.6) |  |
| 30-44 | 44,698 (30.8) | 3,212 (33.9) | 47,910 (31.0) |  |
| 45-59 | 29,892 (20.6) | 1,822 (19.3) | 31,714 (20.5) |  |
| 60 and above | 17,170 (11.8) | 613 (6.5) | 17,783 (11.5) |  |
| Self-reported Race/ Ethnicity |  |  |  |  |
| White | 55,515 (46.5) | 2,603 (43.3) | 58,118 (46.3) | <0.001 |
| Black/Brown | 62,418 (52.3) | 3,371 (56.0) | 65,789 (52.5) |  |
| Indigenous | 352 (0.3) | 15 (0.2) | 367 (0.3) |  |
| Asian | 1,116 (0.9) | 29 (0.5) | 1,145 (0.9) |  |
| Education<sup>†</sup> |  |  |  |  |
| None | 9,062 (8.8) | 717 (9.7) | 9,779 (8.9) | <0.001 |
| ≤7 years | 77,851 (75.5) | 6,004 (81.1) | 83,855 (75.9) |  |
| 8 – 11 years | 11,272 (10.9) | 468 (6.3) | 11,740 (10.6) |  |
| 12 or more years | 4,862 (4.7) | 215 (2.9) | 5,077 (4.6) |  |
| *Clinical conditions and tuberculosis characteristics* |  |  |  |  |
| HIV | 8,345 (6.2) | 1,064 (12.0) | 9,409 (6.5) | <0.001 |
| AIDS among (PLHIV) | 7,442 (89.2) | 930 (87.4) | 8,372 (89.0) | 0.082 |
| Diabetes | 10,256 (7.1) | 485 (5.1) | 10,741 (6.9) | <0.001 |
| Mental health condition | 2,116 (1.5) | 140 (1.5) | 2,256 (1.5) | 0.868 |
| Other immunosuppressive conditions than HIV | 1,667 (1.1) | 82 (0.9) | 1,749 (1.1%) | 0.012 |
| Clinical form |  |  |  |  |
| Pulmonary | 118,277 (81.5) | 8,516 (90.0) | 126,793 (82.0) | <0.001 |
| Extrapulmonary | 22,706 (15.6) | 689 (7.3) | 23,395 (15.1) |  |
| Pulmonary and extrapulmonary | 4,131 (2.8) | 259 (2.7) | 4,390 (2.8) |  |
| Mode of disease discovery |  |  |  |  |
| Outpatient visit | 75,869 (53.6) | 4,693 (50.8) | 80,562 (53.5) | <0.001 |
| Diagnosis clarification during hospitalization | 22,624 (16.0) | 1,261 (13.7) | 23,885 (15.8) |  |
| Active search in institution | 7,606 (5.4) | 900 (9.7) | 8,506 (5.6) |  |
| Emergency / Urgent Visit | 27,005 (19.1) | 1,877 (20.3) | 28,882 (19.2) |  |
| Active community search | 3,364 (2.4) | 270 (2.9) | 3,634 (2.4) |  |
| Contact investigation | 5,018 (3.5) | 230 (2.5) | 5,248 (3.5) |  |
| Laboratory confirmation | 111,993 (77.2) | 8,266 (87.3) | 120,259 (77.8) | <0.001 |
| Sputum culture result |  |  |  |  |
| Positive | 46,200 (31.8) | 4,182 (44.2) | 50,382 (32.6) | <0.001 |
| Negative | 17,727 (12.2) | 1,031 (10.9) | 18,758 (12.1) |  |
| Not performed / No information | 81,188 (55.9) | 4,251 (44.9) | 85,439 (55.3) |  |
| Treatment administration type |  |  |  |  |
| Supervised | 109,710 (81.2) | 7,527 (85.0) | 117,237 (81.4) | <0.001 |
| Self-Administered | 25,392 (18.8) | 1,331 (15.0) | 26,723 (18.6) |  |
| Supervised treatment effectiveness |  |  |  |  |
| Effective | 65,591 (59.8) | 4,503 (59.8) | 70,094 (59.8) | 0.923 |
| Hospitalization | 27,925 (19.2) | 3,170 (33.5) | 31,095 (20.1) | <0.001 |
| *Substance-related behavioral risk factors<sup>†</sup>* |  |  |  |  |
| Alcohol use | 21,630 (15.4) | 2,008 (21.5) | 23,638 (15.8) | <0.001 |
| Illicit drug use | 19,748 (14.1) | 2,199 (23.6) | 21,947 (14.7) | <0.001 |
| Tobacco use | 30,771 (22.0) | 2,466 (26.4) | 33,237 (22.3) | <0.001 |
| *Social vulnerability<sup>†</sup>* |  |  |  |  |
| Incarcerated | 17,594 (12.6) | 2,480 (26.6) | 20,074 (13.4) | <0.001 |
| Homelessness | 2,881 (2.1) | 486 (5.2) | 3,367 (2.3) | <0.001 |

Note: Descriptive statistics are presented as N (%) for categorical variables and mean (standard deviation) for continuous variables. All variables were collected at the time of the first tuberculosis episode. Comparisons between individuals with and without recurrence were performed using Pearson's chi-squared test for categorical variables and Student's t-test for continuous variables. P-values <0.05 were considered statistically significant. Percentages of missing data were race (18.9%), education (24.0%), HIV status (6.5%), treatment administration type (7.1%), and supervised treatment effectiveness (24.3%). † Variable available only for participants aged ≥15 years (n=149,356).

Participants contributed 839,532.6 person-years of follow-up. A total of 9,464 first tuberculosis recurrence events were recorded during a median follow-up time of 5.4 years (IQR: 2.5–8.2). There were 128 recurrences among 5,223 survivors <15 years and 9,336 among 149,356 survivors ≥15 years. The incidence rate of tuberculosis recurrence was 1,113 per 100,000 person-years. When stratified by age group, the incidence rate was 1,153 per 100,000 person-years among individuals aged ≥15 years and 432 per 100,000 person-years among those aged <15 years. The incidence of recurrence peaked at 1,909.4 per 100,000 between 3 and 12 months after treatment completion, remaining above 1,000 per 100,000 person-years between two and five years before declining progressively. More than half (561,531; 54.5%) of the recurrences occurred after 2 years (Table 2).

**Table 2.** Time to tuberculosis recurrence among the analytic cohort for first tuberculosis recurrence, São Paulo, Brazil, 2013 – 2024 (n = 9,464)

| Time to recurrence (interval) | PY | Recurrences (n) | Rate (/100,000 PY) | % of recurrences |
| --- | --- | --- | --- | --- |
| ≤3 months | 37,819.1 | 501 | 1,325.2 | 5.3 |
| 3–12 months | 109,414.6 | 2,090 | 1,909.4 | 22.1 |
| 1–2 years | 130,767.4 | 1,719 | 1,314.8 | 18.2 |
| 2–5 years | 308,104.8 | 3,209 | 1,041.6 | 33.9 |
| >5 years | 253,426.7 | 1,945 | 767.5 | 20.6 |
| Total | 839,532.6 | 9,464 | 1,127.2 | 100.0 |

Note: Time to recurrence was calculated from the end date of the first tuberculosis episode treatment to the start date of the subsequent episode treatment. PY = person-years at risk accrued within each time window among individuals who remained alive and without a first tuberculosis recurrence up to that time. Rate (/100,000 PY) = incidence rate of first tuberculosis recurrence per 100,000 person-years in that interval. % of all recurrences = proportion of the 9,464 first recurrences that occurred in that time window, considering the entire cohort.

Of 2,274 individuals who died after completing tuberculosis treatment and before a recurrent tuberculosis diagnosis, the mean age at their initial tuberculosis episode was 54.0 years (SD 15.9). Median time to death was 2.6 years (IQR: 1.2–4.7). In this group, 21% of deaths occurred within 12 months and 42% within 24 months of treatment completion.

Individuals who experienced tuberculosis recurrence were more often male and younger, with a higher proportion self-identifying as Black or Brown, and the majority had fewer than eight years of education. Incarceration and homelessness during the first tuberculosis episode were also more frequent in this group. HIV was more common among individuals with recurrence, while diabetes was slightly less frequent. Behavioral risk factors, including alcohol use, illicit drug use, and tobacco use, were more common among individuals who had recurrence than those who did not. Pulmonary tuberculosis, positive sputum culture, and laboratory confirmation were more frequently reported, as was hospitalization during the first episode (Table 1).

In the overall population, by the end of the study follow-up period, the cumulative incidence of tuberculosis recurrence was 9.9%, accounting for death as a competing risk (Figure 2). Among people living with HIV (PLHIV), the 12-year cumulative incidence of recurrence reached 17.5%, compared to 9.3% among HIV-negative individuals (p < 0.001) (Figure 2).

**Figure 2.**
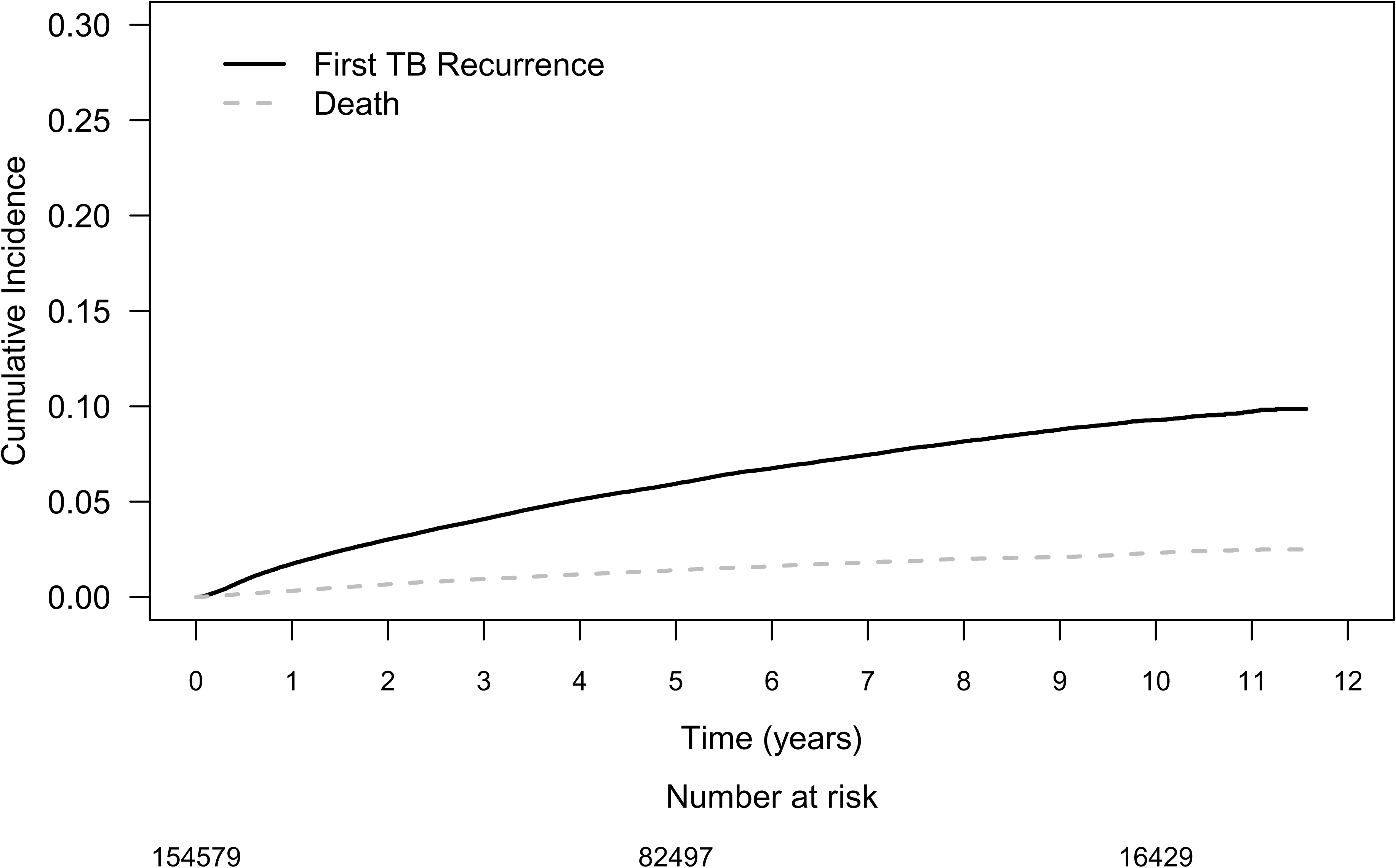
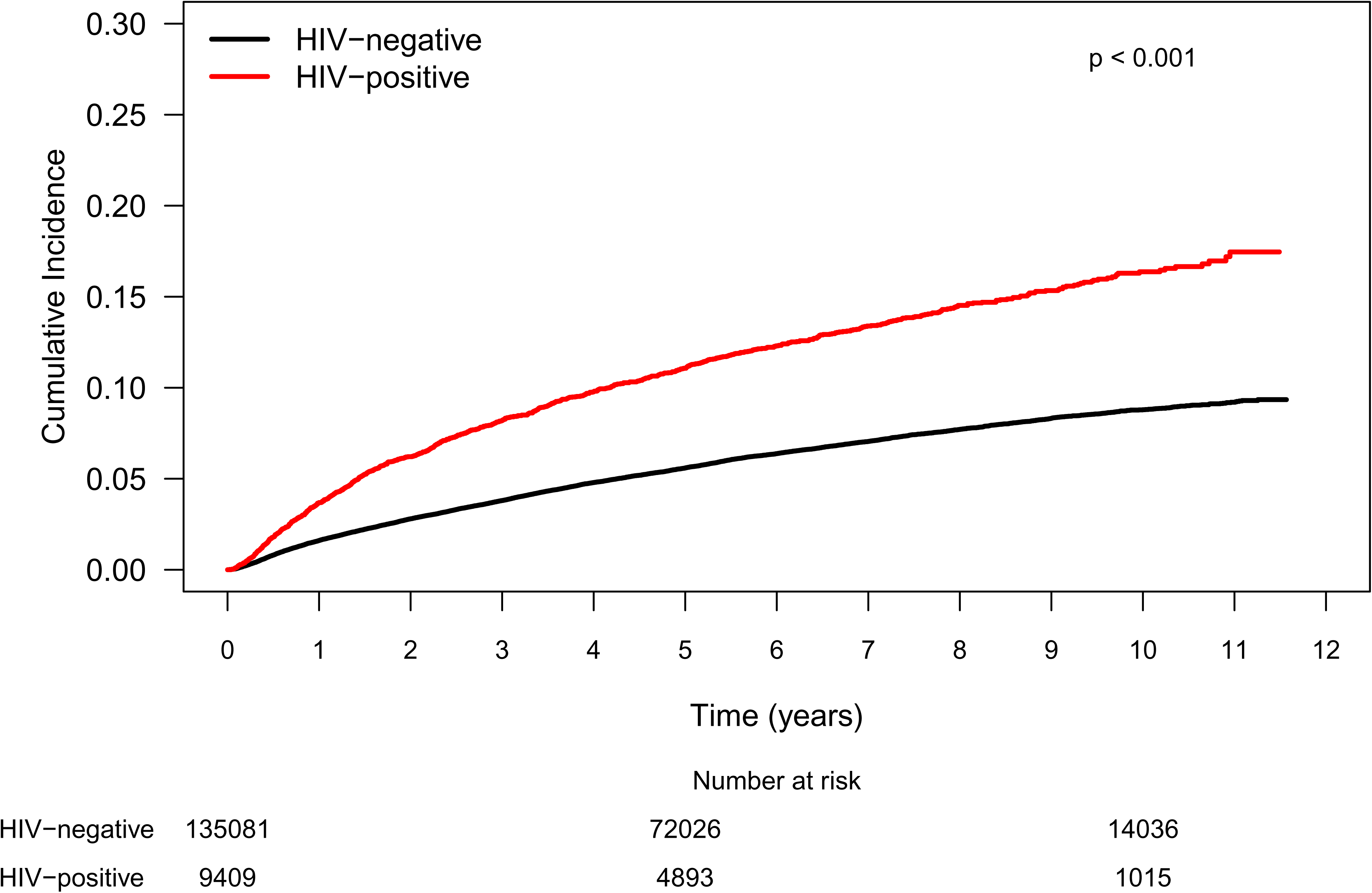
Cumulative incidence curves (CIF) of first tuberculosis recurrence, overall and by HIV status, with death as a competing risk. **(N** = **154,579)** (A) CIF curves for tuberculosis recurrence and death as a competing risk in the study population. (B) CIF curves for tuberculosis recurrence stratified by HIV status. Group comparisons were evaluated using Gray’s test. "Number at risk" indicates the number of individuals still at risk at 0, 5, and 10 years of follow-up.

Higher 12-year cumulative recurrence rates were also observed among individuals reporting illicit drug use (18.0% vs. 8.9%, p < 0.001), alcohol use (13.6% vs. 9.4%, p < 0.001), and among those experienced homelessness (24.4% vs. 9.8%, p < 0.001), and incarceration (19.7% vs. 8.4%, p < 0.001) during their first treatment episode (Figure 3).

**Figure 3.**
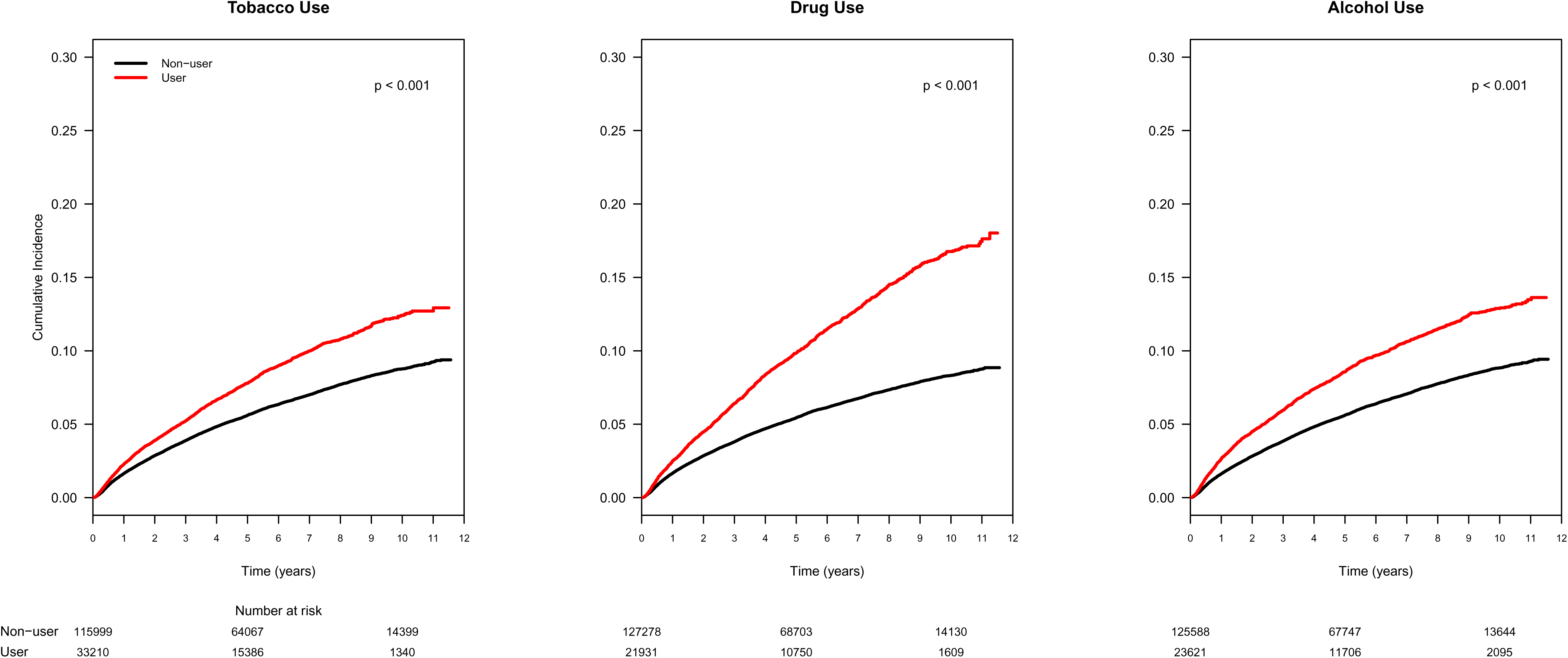
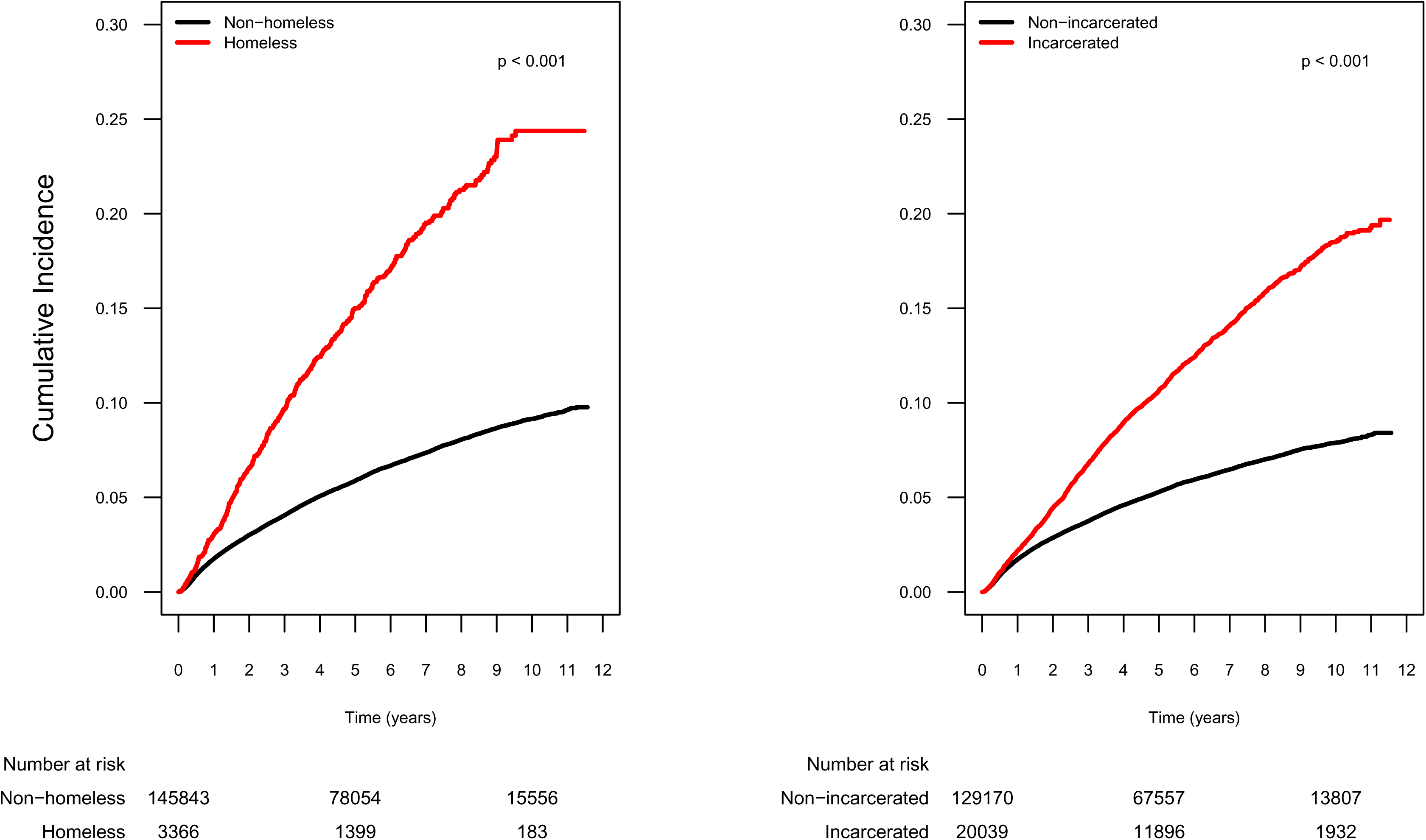
Cumulative incidence curves (CIF) of tuberculosis recurrence by substance use and social vulnerability among individuals aged. ≥**15 years (n=149,356)** (A) CIF curves for tuberculosis recurrence stratified by substance use behaviors, including tobacco use, illicit drug use, and alcohol use. (B) CIF curves are stratified by social vulnerability indicators, including homelessness and incarceration experience during the first tuberculosis episode. Group comparisons were evaluated using Gray’s test. "Number at risk" indicates the number of individuals still at risk at 0, 5, and 10 years of follow-up.

In the multivariable competing-risk model (Table 3), male sex (aSHR 1.25, 95% CI 1.15–1.35) and younger age were associated with recurrence. Recurrence risk decreased with higher education: 0.64 (0.55–0.75) for 8–11 years and 0.52 (0.41–0.66) for ≥12 years vs none, showing a dose–response gradient. Race was not associated with recurrence, while pulmonary tuberculosis (2.10, 1.87–2.35), HIV (1.69, 1.54–1.86), and hospitalization (2.53, 2.36–2.71) were strong clinical predictors. No difference in recurrence was seen between self-administered and directly observed therapy. Behavioral risk factors such as alcohol use (aSHR 1.14, 95% CI 1.05–1.25) and illicit drug use (aSHR 1.22, 95% CI 1.12–1.32) were also significantly associated with higher recurrence risk, though tobacco use was not. Markedly elevated risks were observed among individuals experiencing social vulnerability. Individuals who experienced homelessness or incarceration during the first tuberculosis episode had markedly higher recurrence risks (aSHR 1.65, 95% CI 1.40–1.95; and 2.30, 95% CI 2.13–2.48, respectively).

**Table 3.** Subdistribution hazard ratios (SHR) for first tuberculosis recurrence among individuals aged. ≥**15 years, based on complete-case analysis using competing-risks regression with death as a competing event**

| Variable | Unadjusted SHR<br>(95% CI) | Adjusted SHR<br>(95% CI) |
| --- | --- | --- |
| *Demographic* |  |  |
| Sex: male | 1.77 (1.68–1.86) | 1.25 (1.15–1.35) |
| Age (ref: 15–29 years) |  |  |
| 30–44 years | 0.97 (0.92–1.02) | 0.89 (0.83–0.95) |
| 45–59 years | 0.83 (0.79–0.88) | 0.91 (0.83–0.99) |
| ≥60 years | 0.51 (0.47–0.55) | 0.69 (0.60–0.79) |
| Race (ref: White) |  |  |
| Black or Brown | 1.19 (1.13–1.25) | 1.04 (0.98–1.11) |
| Indigenous and Asian | 0.92 (0.56–1.54) | 0.92 (0.66–1.28) |
| Education (ref: none) |  |  |
| Education: ≤7 years | 1.01 (0.94–1.10) | 0.88 (0.80–0.97) |
| Education: 8–11 years | 0.61 (0.54–0.68) | 0.64 (0.55–0.75) |
| Education: ≥12 years | 0.45 (0.38–0.53) | 0.52 (0.41–0.66) |
| *Clinical conditions and tuberculosis characteristics* |  |  |
| HIV | 1.97 (1.85–2.10) | 1.69 (1.54–1.86) |
| Diabetes | 0.75 (0.68–0.82) | 0.91 (0.79–1.05) |
| Mental health condition | 0.87 (0.73–1.04) | 0.95 (0.73–1.26) |
| Other immunosuppression condition than HIV | 0.78 (0.71–0.82) | 0.87 (0.61–1.23) |
| Tuberculosis anatomic classification<br>(ref: extrapulmonary) |  |  |
| Pulmonary | 2.30 (2.12–2.49) | 2.10 (1.87–2.35) |
| Pulmonary + Extrapulmonary | 2.04 (1.76–2.36) | 1.28 (1.04–1.58) |
| Hospitalization | 2.06 (1.97–2.15) | 2.53 (2.36–2.71) |
| Self-administered treatment (ref: supervised) | 0.78 (0.73–0.82) | 1.02 (0.94–1.12) |
| *Substance-related behavioral risk factors* |  |  |
| Alcohol use | 1.53 (1.46–1.61) | 1.14 (1.05–1.25) |
| Illicit drug use | 1.93 (1.84–2.02) | 1.22 (1.12–1.32) |
| Tobacco use | 1.42 (1.35–1.48) | 0.99 (0.92–1.07) |
| *Social vulnerability* |  |  |
| Incarcerated | 2.32 (2.22–2.43) | 2.30 (2.13–2.48) |
| Homelessness | 3.17 (2.90–3.48) | 1.65 (1.40–1.95) |

Note: SHR: subdistribution hazard ratio from Fine and Gray competing risks model. CI = Confidence Interval. Reference categories are indicated in parentheses. All variables were collected during the first tuberculosis episode. All estimates were adjusted for all other variables listed in the table. The final model included 90,123 individuals, 4,552 recurrence events, and 1,433 competing events. Collinearity was assessed using Pearson correlation and variance inflation factors (VIF) with a cutoff of 5; no substantial collinearity was detected.

In sensitivity analysis, SHRs were consistent between the imputed and complete-case models (Appendix Table 3).

Among children (Table 4), male sex was associated with lower recurrence risk (aSHR 0.53, 95% CI 0.31–0.91), while risk increased 16% per additional year of age (1.16, 1.09–1.25).

**Table 4.** Subdistribution hazard ratios (SHR) for first tuberculosis recurrence in individuals aged <15 years (complete-case analysis)

| Variable | Unadjusted SHR<br>(95% CI) | Adjusted SHR<br>(95% CI) |
| --- | --- | --- |
| Sex: male | 0.62 (0.43–0.88) | 0.53 (0.31–0.91) |
| Age (per year increase) | 1.14 (1.09–1.19) | 1.16 (1.09–1.25) |
| Race: non-White<br>(Ref: White) | 0.96 (0.60–1.52) | 0.84 (0.51–1.40) |
| Pulmonary form<br>(Ref: any extrapulmonary form) | 2.21 (1.34–3.65) | 2.60 (1.25–5.38) |
| HIV | 7.00 (3.39–14.46) | 6.38 (2.64–15.40) |
| Hospitalization | 1.91 (1.34–2.72) | 2.82 (1.65–4.80) |

Note: Age was modeled as a continuous variable. Reference categories are indicated in parentheses. SHR = subdistribution hazard ratio estimated from a Fine and Gray competing risks regression model. All variables were collected during the first tuberculosis episode. The final model for individuals aged <15 years included 3,333 cases, 63 recurrence events, 2 competing events, and 3,268 censored cases.

HIV (6.38, 2.64–15.40) and hospitalization during the first episode (2.82, 1.65–4.80) were the strongest predictors of recurrence. Pulmonary tuberculosis was associated with significantly higher risk of recurrence compared to extrapulmonary disease (aSHR 2.60, 95% CI 1.25–5.38, p = 0.010). Race was not associated with recurrence.

Results were consistent in sensitivity analyses (Appendix Table 4).

## Discussion

In a large cohort of patients who completed treatment for tuberculosis, we identified very high rates of recurrence, 1,127.2 per 100,000 person-years, comparable to the incidence of new tuberculosis among people living with HIV in Brazil (1,000 per 100,000). ^12^ The risk of recurrence was highest in the first year after treatment completion, particularly between 3 and 12 months (1,909.4 per 100,000 person-years). However, more than half of all recurrent episodes (54.5%) occurred after two years, including 33.9% between two and five years after treatment completion, when recurrence rates remained above 1,000 per 100,000 person-years before declining thereafter. Higher recurrence risk was associated with demographic factors (male sex, low education level), clinical conditions (HIV infection, pulmonary tuberculosis, hospitalization), behavioral factors (alcohol use, drug use), and social vulnerabilities (homelessness and incarceration). Together, these results underscore the need for sustained monitoring of tuberculosis survivors for recurrence for at least five years post-treatment, particularly those with biomedical and social vulnerabilities.

Our study could not differentiate relapse from reinfection. However, the literature suggests that early recurrences are often related to host or treatment factors (relapse), whereas later events are more likely to represent reinfection in high-burden settings. ^15–17^ In our study, approximately half of recurrent episodes occurred within two years, and half occurred thereafter, reflecting the multiple determinants and mechanisms influencing recurrence risk.

HIV is a well-established risk factor for tuberculosis recurrence, particularly in high-burden settings ^7^. Our findings reflect this association and extend it to children, an underrepresented population in recurrence research. ^18^ We did not identify a significant association between diabetes and tuberculosis recurrence, though prior studies have shown that diabetes may increase the risk of recurrence. ^19^ Although diabetes screening is recommended, it is not mandatory under Brazilian tuberculosis guidelines. The recommendation was only introduced in 2019, ^13^ which likely contributed to underdiagnosis and exposure misclassification.

Lower recurrence rates in older individuals likely reflect the competing risk of death from other causes rather than a true protective effect of age. Although previous studies have reported higher recurrence among older adults ^16,20^, many did not model death as a competing event.

When death is censored rather than treated as a competing event, the cumulative incidence of recurrence tends to be overestimated, with bias amplified in subgroups with higher mortality, such as older adults. ^14^ Competing-risk methods provide estimates of recurrence probabilities that appropriately account for differential mortality across age groups.

Recurrence was more common among adult men, consistent with previous studies. ^21–23^ This pattern may reflect men’s greater exposure to high-risk environments, including male-dominated workplaces (e.g., manufacturing, transportation), informal labor markets with poor ventilation, overcrowded urban housing, and social venues such as bars or shelters, often coupled with higher prevalence of behavioral risk factors such as alcohol, tobacco, and drug use. ^22^

In our cohort, alcohol and illicit drug use were associated with an increased risk of recurrence, while tobacco use was not after covariate adjustment. Although smoking has been linked to worse tuberculosis outcomes and higher recurrence risk, ^24^ the absence of an association in our study may reflect limited exposure data, as information on smoking duration and intensity were not available, and smoking overlaps with other behavioral and social vulnerabilities.

Incarceration during the first tuberculosis episode was one of the strongest predictors of recurrence, with a greater effect size than HIV. Prisons in Latin America have been described as key tuberculosis reservoirs, reflecting concentrated transmission in settings with limited infection control strategies. ^25,26^ In Brazil, a recent modeling study estimated 36.9% of incident tuberculosis cases were attributable to incarceration, more than to HIV, alcohol use, or undernutrition. ^25^ Considering its long-term influence on exposure and vulnerability, incarceration history should be accounted for in post-treatment care and recurrence risk assessments.

In our cohort, individuals who experienced homelessness during their first tuberculosis episode had higher recurrence risk. This population is frequently studied for delayed diagnosis and poor treatment adherence, yet few investigations have explicitly examined post-treatment outcomes. A systematic review identified housing affordability and stability as critical determinants across the tuberculosis care continuum, including recurrence risk. ^27^ People experiencing homelessness often face overlapping vulnerabilities — substance use, malnutrition, and limited access to care—that may disrupt treatment continuity and follow-up. ^27,28^ They also remain exposed to high-transmission environments, including overcrowded and poorly ventilated shelters, which may increase risk of tuberculosis re-exposure. ^28^

Hospitalization during the first tuberculosis episode was identified as a strong predictor of recurrence. This association may reflect persistent individual vulnerability, such as underlying immunological or biological susceptibility not captured by available variables ^29^, or structural vulnerability, with hospitalization serving as a proxy for barriers to timely diagnosis and treatment that allow disease progression and complications. ^30^

We found no association between supervised treatment and recurrence risk, likely because supervision primarily benefits treatment completion. Since our cohort included only individuals who successfully completed treatment, the protective effect of supervision was likely embedded in the inclusion criteria, limiting its ability to differentiate recurrence risk.

Tuberculosis recurrence demonstrates that disease risk persists beyond treatment completion, as survivors remain exposed to the same biological and social determinants of their initial disease. Providing screening and treatment for non-tuberculosis conditions during or after treatment for tuberculosis would reduce risk for future tuberculosis episodes and other tuberculosis sequelae. At a minimum, differentiated post-treatment follow-up is needed, with structured surveillance incorporated into national guidelines—particularly for individuals with clinical and/or social vulnerabilities. Integration with social and health services is crucial to reduce the long-term burden and prevent delayed detection of recurrent disease.

These findings should be interpreted in light of several limitations. Although we had access to treatment duration and follow-up smear microscopy results, smear data were not available for all cases, and no variables captured whether clinical or radiological criteria were applied, particularly relevant for extrapulmonary or pediatric tuberculosis. Sputum culture results were unavailable, preventing the application of WHO definitions for cure in drug-resistant tuberculosis. ^4^ Resistance data were also incomplete, though the overall prevalence of drug-resistant TB in São Paulo is low (1.1%). ^12^

Behavioral variables like tobacco use, alcohol consumption, and illicit drug use, along with social vulnerability indicators such as homelessness and incarceration, were recorded only at baseline. We lacked information on duration, recurrence, or continuity of these exposures, which limited our ability to assess their cumulative impact or changes in exposure patterns during follow-up. Additionally, our dataset had limited detailed clinical comorbidity information, such as nutritional status, diabetes care, and antiretroviral therapy (ART) use among PLHIV. Other comorbidities that may have influenced treatment choices or risk of complications during the disease could not be evaluated.

Tuberculosis recurrence may have been underestimated due to the geographic scope of the notification system, which only captures tuberculosis cases reported within São Paulo state. Consequently, recurrences occurring in other states were not captured. In contrast, probabilistic linkage with the national mortality database enabled identification of post-treatment deaths nationwide. However, the linkage process had limited sensitivity and may have missed some deaths, leading to underestimation of competing mortality risk.

Our findings reinforce that people who complete tuberculosis treatment remain at elevated risk of recurrence. The peak in recurrence risk within the first year marks a critical window during which structured follow-up strategies may have the greatest impact, while the persistence of elevated rates through the five years suggests that surveillance should not be discontinued prematurely. Yet, the current global recommendation for post-treatment monitoring is only conditional due to weak evidence. Incorporating targeted follow-up approaches for individuals with greater clinical and social vulnerability may facilitate earlier detection, identify opportunities for preventing recurrence, and support a broader view of the tuberculosis care continuum that extends beyond treatment completion.

## Data share statement

The data used in this study were obtained from the São Paulo State Tuberculosis Program (TBweb) and linked with the Brazilian Mortality Information System (SIM). These administrative health databases are not publicly available due to confidentiality restrictions. Data access requires formal authorization from the São Paulo State Department of Health.

## Contributors

E. Lepka de Lima, J.E. Golub, and S.R. Cox conceived and designed the study. A.A. Lindoso and S. Fukasava acquired and curated the data. E. Lepka de Lima, I. Salles, and S. Cohn performed the data analysis. All authors interpreted the results. E. Lepka de Lima wrote the first draft of the manuscript. J.E. Golub, S.R. Cox, and I. Salles revised the manuscript with important intellectual contributions. All authors read and approved the final manuscript.

## Data Availability

The data used in this study come from the Sao Paulo State TB surveillance system (TBweb) and the Brazilian Mortality Information System (SIM). Due to privacy and data protection regulations, these datasets cannot be shared publicly. De-identified data may be made available upon reasonable request and with authorization from the Sao Paulo State Epidemiologic Surveillance Center.

## Acknowledgments

We thank the Tuberculosis Surveillance Team of São Paulo State and the Epidemiological Surveillance Division for their support with data access and linkage procedures.

