## Supplementary material for "Risk and determinants of tuberculosis recurrence: a 12-year population-based cohort study"

**1. Variable dictionary**
**2. Linkage methods**
**3. Supplementary tables**

**Table S1.** Detailed exclusion criteria and number of cases removed at each stage of data processing and cohort definition.
**Table S2.** Standardized treatment outcome definitions – WHO framework.
**Table S3**. Subdistribution hazard ratios (SHRs) for first tuberculosis recurrence among individuals aged ≥15 years, based on imputed data.
**Table S4.** Subdistribution hazard ratios (SHRs) for first tuberculosis recurrence among individuals aged <15 years, based on imputed data.

### 1. Variable Dictionary

*A. Identification and demographics*

1. SINAN number: A unique patient identifier assigned to each patient, ensuring that each case is tracked accurately.
2. Race: The patient's race, categorized according to system-defined options.
3. Country of origin: The country where the patient was born.
4. Date of birth: The patient's date of birth, formatted as day/month/year.
5. Age at diagnosis: The system automatically calculates the patient's age at the time of tuberculosis diagnosis.
6. Sex: The patient's sex at birth, classified as male or female.
7. Education level: The patient's highest level of education, based on years of schooling.
8. Occupation: The patient's occupation, categorized according to system options.

*B. Diagnosis and treatment*

9. Treatment sequence: The number of treatment courses the patient has undergone, indicating the treatment regimen's sequence.
10. Notification date: The date the case was reported to the system.
11. Treatment start date: The date when the patient’s treatment for tuberculosis began.
12. Diagnosis date: The date when the patient was officially diagnosed with tuberculosis.
13. Case type: The type of case may include "New," "Recurrence," "Abandonment," and other categories.
14. Case outcome: The outcome of the tuberculosis case, including outcomes such as "Cured," "Death from tuberculosis," and "Death from other causes."
15. Treatment end date: When the patient’s treatment was completed or ended.

*C. Clinical forms and classification*

16. Primary clinical form: The primary form of tuberculosis could be pulmonary or extrapulmonary.
17. Secondary clinical form: The secondary form of tuberculosis, if applicable.
18. Tertiary clinical form: The tertiary form of tuberculosis, if applicable.
19. Anatomical clinical classification: The classification of tuberculosis as pulmonary, extrapulmonary, or mixed/disseminated forms.

*D. Laboratory tests and diagnosis*

20. Laboratory confirmation: Whether laboratory tests, such as sputum smear microscopy, molecular testing, or culture, confirm the tuberculosis diagnosis.
21. Sputum culture: The result of sputum culture tests to detect tuberculosis bacilli.
22. Other site culture: Culture tests performed on biological samples other than sputum, when applicable.

*E. Clinical and social conditions*

23. HIV status: The patient’s HIV status, indicating whether they are HIV positive or negative.
24. AIDS status: Whether the patient has been diagnosed with AIDS.
25. Diabetes status: Whether the patient was reported to have diabetes, as recorded by the health professional at notification (based on medical diagnosis or self-reported history).
26. Alcoholism status: Whether the patient has a history of alcoholism.
27. Mental health conditions: Whether the patient has any diagnosed mental health issues.
28. Drug use history: Whether the patient has a history of substance use.

29. Other immunosuppressive conditions: Any other conditions besides HIV/AIDS that may compromise the immune system.
30. Tobacco use history: Whether the patient has a history of tobacco use.

F. *Contacts and address*

31. Total contacts: The number of people the patient was in contact with during the contagious period.
32. Examined contacts: The number of contacts examined to check for possible tuberculosis transmission.
33. Contact disease: The disease identified in the patient's contacts, if any.

G. *Treatment and management*

34. Address type: The type of patient address is categorized as fixed residence, no fixed residence (homeless), or detainee.
35. Treatment city: The city where the patient is receiving treatment.
36. Treatment administration type: Whether the patient's treatment is supervised or not.
37. Supervised treatment effectiveness: variable indicating whether directly observed treatment (DOT) was conducted effectively, defined as at least three supervised doses per week during treatment.

H. *Resistance and bacteriological tests*

38. Bacteriological test results: Sputum smear microscopy results performed from the 1st to the 9th month of treatment were categorized as positive, negative, or not performed (e.g., missing information, ongoing, or not performed).
39. Molecular test: The result of molecular testing, indicating whether the GeneXpert® rapid molecular test was performed.

I. *Cause of death and resistance*

40. Date of death: The date of the patient's death, if applicable.
41. Cause of death: The cause of death, described in Brazilian code (CID 10), whether it was due to tuberculosis or another condition.
42. Drug resistance: The patient's resistance status to the tuberculosis drugs, if identified.
43. Rifampicin sensitivity: The patient's sensitivity to rifampicin if it was performed.
44. Isoniazid sensitivity: The patient's sensitivity to isoniazid if it was performed.

J. *Additional Information*

45. Disease discovery: The method by which the tuberculosis disease was discovered, which could include various options such as outpatient demand, diagnosis during hospitalization, active search in institutions, and others.
46. Hospital admission: Indicates whether the patient was admitted to the hospital for treatment, categorizing admission into different types, such as emergency, elective, or others.

### 2. Linkage methods

A probabilistic record linkage was conducted between tuberculosis cases reported in São Paulo from 2013 to 2024 and death records from Brazil's Mortality Information System (SIM) for the same period to ascertain mortality after tuberculosis notification. Data from TBweb (n = 249,208) and SIM were imported into Microsoft Access (Microsoft Corp., Redmond, USA). The matching process used key identifiers: the patient's name, the mother's name, and the date of birth. Microsoft Access's matching features were used to compute similarity scores between record pairs across both datasets. Blocking strategies were applied to optimize comparisons and reduce false positives.

The automated linkage identified an initial set of 12,813 potential matches. A manual review followed, in which record pairs with discrepancies, incomplete fields, or non-informative values (e.g., "unknown," "ignored") were excluded. After validation, 10,056 record pairs were retained as high-confidence matches between tuberculosis notifications and corresponding deaths.

After the linkage step, all personally identifiable information was removed to ensure data confidentiality and compliance with ethical and data protection standards.

**3. Supplementary tables**

**Table S1. Detailed exclusion criteria and the number of cases removed at each stage of data processing and cohort definition.**

| Structural Category | Exclusion Criterion | Technical Justification | n |
|---|---|---|---|
| 1. Not at risk for first tuberculosis recurrence | Diagnosis reclassified as non-tuberculosis | Case does not meet tuberculosis definition; excluded from target population. | 8,315 |
| | Death during the first tuberculosis episode. | Patient did not survive to achieve cure; no risk of recurrence. | 17,695 |
| | Notifications with outcomes other than ‘cure’, such as abandonment, failure, treatment change, missed follow-up, or still recorded as “on treatment” (ambulatory or hospitalized). | Outcome known, but not consistent with cured status; no valid baseline for follow-up. | 27,754 |
| Subtotal (1) | | | 53,764 |
| 2. No evaluable due incomplete information | Transferred out of the state of São Paulo during the first episode. | Final outcome not observable within surveillance system. | 1,861 |
| | Undefined or missing treatment outcome in the first tuberculosis episode. | Outcome not recorded or classified; impossible to determine ‘cure’ status. | 12,481 |
| | Irreparable inconsistencies or duplicate records. | Data quality issues prevent inclusion. | 38 |
| Subtotal (2) | | | 14,380 |
| 3. Invalid treatment timeline | Missing cure date or zero-day interval between cure and recurrence. | Cannot define follow-up start or confirm that recurrence is a new event. | 858 |
| | Recurrence recorded on the same day as cure (residual cases). | Ambiguity between continuation vs. true new episode. | 3 |
| | Cure date after December 30, 2024. | No available follow-up time within study window. | 3 |
| | Treatment duration <6 months (or <5 months without smear conversion at months 5–6) | Does not meet treatment duration criteria to ensure treatment adequacy. | 4,386 |
| | Treatment duration >2·5 years | Does not meet treatment duration criteria to ensure treatment adequacy. | 1,170 |
| Subtotal (3) | | | 6,420 |
| 4. Subsequent notifications not eligible for time-to-first recurrence analysis | Retreatment episodes or multiple recurrences in the same individual | Only the first recurrence was analyzed; additional episodes excluded to prevent duplication. | 20,765 |
| Subtotal (4) | | | 20,765 |
| Total excluded | | | 95,329 |
| Final analytical cohort | | | 154,579 |

**Table S2. Standardized Treatment Outcome Definitions – WHO Framework**

**A.2.1. Outcomes for Drug-Susceptible Tuberculosis**

| Outcome | Definition |
|---|---|
| Cured | A pulmonary tuberculosis patient with bacteriologically confirmed TB at the beginning of treatment who was smear- or culture-negative in the last month and on one prior occasion. |
| Treatment completed | A TB patient who completed treatment without evidence of failure, but with no record of smear or culture results in the last month and prior occasion. |
| Treatment failed | A TB patient whose sputum smear or culture is positive at month 5 or later during treatment. |
| Died | A TB patient who dies for any reason before starting or during the course of treatment. |
| Lost to follow-up | A TB patient who did not start treatment or whose treatment was interrupted for 2 consecutive months or more. |
| Not evaluated | A TB patient for whom no treatment outcome is assigned, including "transferred out" and cases with unknown outcome. |
| Treatment success | The sum of cured and treatment completed. |

Note: Patients found to have rifampicin-resistant (RR), multidrug-resistant (MDR), or extensively drug-resistant (XDR) TB at any point are excluded from this cohort and instead included in A.2.2.

**A.2.2. Outcomes for RR-TB / MDR-TB / XDR-TB**

| Outcome | Definition |
|---|---|
| Cured | Treatment completed as per national policy without evidence of failure, and ≥3 consecutive negative cultures taken ≥30 days apart after the intensive phase. |
| Treatment completed | Treatment completed per national policy without evidence of failure, but without record of ≥3 negative cultures post-intensive phase. |
| Treatment failed | Treatment terminated or major regimen change (≥2 drugs) due to:<br>– Lack of conversion by end of intensive phase<br>– Reversion to positive in continuation phase<br>– Acquired resistance to fluoroquinolones or second-line injectables<br>– Severe adverse drug reactions (ADRs). |
| Died | Patient who dies for any reason during treatment. |
| Lost to follow-up | Treatment interrupted for ≥2 consecutive months. |
| Not evaluated | No assigned outcome (includes "transferred out" and unknown outcomes). |
| Treatment success | The sum of cured and treatment completed. |

**Table S3. Subdistribution hazard ratios (SHRs) for first tuberculosis recurrence among individuals aged ≥15 years, based on imputed data. Estimates were derived using competing-risks regression with death as a competing event**

| Variable | Unadjusted SHR (95% CI) | Adjusted SHR (95% CI) |
|---|---|---|
| *Demographic* | | |
| Sex: male | 1·77 (1·68–1·86) | 1·26 (1·19–1·33) |
| Age group (ref: 15–29 years) | | |
| 30–44 years | 0·97 (0·92–1·02) | 0·92 (0·88–0·97) |
| 45–59 years | 0·83 (0·79–0·88) | 0·90 (0·85–0·96) |
| ≥60 years | 0·51 (0·47–0·56) | 0·65 (0·60–0·71) |
| Race (ref: White) | | |
| Black or Brown | 1·18 (1·13–1·24) | 1·05 (1·00–1·09) |
| Indigenous and Asian | 0·68 (0·52–0·88) | 0·87 (0·67–1·13) |
| Education (ref: none) | | |
| Education: ≤7 years | 1·01 (0·93–1·09) | 0·98 (0.91–1.07) |
| Education: 8–11 years | 0·66 (0·58–0·74) | 0·79 (0·71–0·89) |
| Education: ≥12 years | 0·54 (0·46–0·64) | 0·69 (0.58–0·82) |
| *Clinical conditions and tuberculosis characteristics* | | |
| HIV | 1·92 (1·80–2·05) | 1·66 (1·56–1·77) |
| Diabetes | 0·75 (0·68–0·82) | 0·96 (0·87–1·06) |
| Mental health condition | 0·87 (0·73–1·04) | 0·85 (0·70–1·01) |
| Other immunosuppressive condition than HIV | 0·78 (0·62–0·97) | 1·00 (0·80–1·24) |
| Tuberculosis anatomic classification (ref: Extrapulmonary) | | |
| Pulmonary | 2·30 (2·13–2·49) | 2·25 (2·07–2·38) |
| Pulmonary + Extrapulmonary | 2·04 (1·67–2·50) | 1·59 (1·40–1·71) |

| | | |
|---|---|---|
| Hospitalization during 1st TB episode | 2·06 (1·97–2·15) | 2.37 (2·26–2·49) |
| Self-administered treatment<br>(ref: supervised) | 0·78 (0·73 – 0·83) | 0·**97** (0.91–1 03) |
| *Substance-related behavioral risk factors* | | |
| Alcohol use | 1·53 (1·46–1·61) | 1·13 (1·06–1·20) |
| Illicit drug use | 1·93 (1·84–2·02) | 1·26 (1·19–1·33) |
| Tobacco use | 1·42 (1·35–1·48) | 1·13 (1·08–1·19) |
| *Social vulnerability* | | |
| Incarcerated | 2·21 (2·11–2·31) | 2·09 (1·98–2·21) |
| Homelessness | 2·67 (2·44–2·92) | 1·78 (1·61–1·96) |

Note: SHR: subdistribution hazard ratio from Fine and Gray competing risks model. CI = Confidence Interval. Reference categories are indicated in parentheses. All variables were collected during the first tuberculosis episode. Analyses were conducted using multiple imputation by chained equations, and parameter estimates, and standard errors were combined across imputed datasets using Rubin's rules. Adjusted estimates were derived from a multivariable model including all covariates listed in the table.

**Table S4. Subdistribution hazard ratios (SHRs) for first tuberculosis recurrence among individuals aged <15 years, based on imputed data. Estimates were derived using competing-risks regression with death as a competing event.**

| Variable | Unadjusted SHR (95% CI) | Adjusted SHR (95% CI) |
|---|---|---|
| *Demographic* | | |
| Sex: male | 0·62 (0·43–0·88) | 0·62 (0·39–0·99) |
| Age (per year increase) | 1·14 (1·09–1·19) | 1·13 (1·07–1·20) |
| Race: non-White (ref: White) | 0·96 (0·60–1·52) | 0·82 (0·52–1·31) |
| *Clinical conditions and TB characteristics* | | |
| Pulmonary form (ref: any extrapulmonary form) | 2·22 (1·35–3·65) | 2·87 (1·43–5·76) |
| HIV | 4·51 (2·23–9·11) | 4·47 (1·89–10·61) |
| Hospitalization | 1·91 (1·34–2·72) | 2·80 (1·68–4·67) |

Note: SHR = subdistribution hazard ratio; CI = confidence interval. All models were estimated using competing-risks regression with death as a competing event, based on imputed data (see Supplementary Table 3 note for details).
